# Clinical course and severity outcome indicators among COVID-19 hospitalized patients in relation to comorbidities distribution: Mexican cohort

**DOI:** 10.1101/2020.07.31.20165480

**Authors:** COVID-19 Hospitalizations in Mexico, Genny Carrillo, Nina Méndez-Domínguez, Kassandra Desire Santos-Zaldivar, Andrea Rochel-Pérez, Mario Azuela-Morales, Osman Cuevas-Koh, Alberto Álvarez-Baeza

## Abstract

**Introduction:** COVID-19 affected worldwide, causing to date, around 500,000 deaths. In Mexico, by April 29, the general case fatality was 6.52%, with 11.1% confirmed case mortality and hospital recovery rate around 72%. Once hospitalized, the odds for recovery and hospital death rates depend mainly on the patients’ comorbidities and age. In Mexico, triage guidelines use algorithms and risk estimation tools for severity assessment and decision-making. The study’s objective is to analyze the underlying conditions of patients hospitalized for COVID-19 in Mexico concerning four severity outcomes.

**Materials and Methods:** Retrospective cohort based on registries of all laboratory-confirmed patients with the COVID-19 infection that required hospitalization in Mexico. Independent variables were comorbidities and clinical manifestations.

**Dependent variables were four possible severity outcomes:** (a) pneumonia, (b) mechanical ventilation (c) intensive care unit, and (d) death; all of them were coded as binary Results: We included 69,334 hospitalizations of laboratory-confirmed and hospitalized patients to June 30, 2020. Patients were 55.29 years, and 62.61% were male. Hospital mortality among patients aged<15 was 9.11%, 51.99% of those aged >65 died. Male gender and increasing age predicted every severity outcome. Diabetes and hypertension predicted every severity outcome significantly. Obesity did not predict mortality, but CKD, respiratory diseases, cardiopathies were significant predictors.

**Conclusion:** Obesity increased the risk for pneumonia, mechanical ventilation, and intensive care admittance, but it was not a predictor of in-hospital death. Patients with respiratory diseases were less prone to develop pneumonia, to receive mechanical ventilation and intensive care unit assistance, but they were at higher risk of in-hospital death.

## Introduction

SARS-CoV-2 is in the family of viruses called coronavirus, characterized for causing mild symptoms like the common cold to severe respiratory syndrome (SR) to susceptible humans [1]. As the cause of an emerging highly transmissible agent, COVID-19 was identified as the cause of severe, fatal respiratory disease in Wuhan China by the end of 2019 [2]. By February 28, imported cases were confirmed in Mexico [3].

COVID-19 has to date, affected worldwide population involving approximately 216 countries, with 11 million confirmed cases and around 500,000 deaths [4]. In Mexico, by April 29, 245,000 confirmed cases, and 29,843 deaths had been reported; based on these data, the national incidence rate was 13.89 / 100,000 inhabitants. General case fatality is 6.52%, and 11.1 % confirmed case mortality and hospital recovery rate around 72%. [3]. Once hospitalized, the odds for recovery and, therefore, the hospital death rates depend mainly on the patients’ underlying health conditions and the clinical-pathological manifestations that they may develop.

According to epidemiologic data, men are more prone to develop severe cases than women are; smoking abuse may relate to adverse outcomes among COVID-19 hospitalized patients [5, 6]. Furthermore, comorbidities have been associated with a higher mortality rate, including hypertension, respiratory diseases, cardiac pathologies, diabetes, and CPK [7-9]. Additionally, smoking habits may relate to adverse outcomes among COVID-19 hospitalized patients due to an overexpression of angiotensin converter enzyme [10]. The mechanical impairment of respiration in COVID-19 patients is due to a nervous alteration associated with damage to the solitary and ambiguous tract nuclei. They are responsible for the innervation of the muscles, glands, and blood vessels, causing death due to dysfunction of the cardiorespiratory system [11,12]. Another cause of mechanical alteration is related to acute respiratory distress syndrome, caused by excessive recruitment of immune cells, producing an increase in pro-inflammatory factors, ending in dysfunction of the pulmonary circulation, and increased cardiac output [13]. Therefore, immune impairment and uncontrolled diabetes may inhibit the proliferation of lymphocytes, monocytes, macrophages, and neutrophils, favoring viral replication, and translating in morbidity that is more significant and mortality proneness [14]. Additionally, patients with long term uncontrolled diabetes may also sum an unknown kidney function impairment.

Obesity is a chronic disease resulting from the storage of energy as fat cells causing hypertrophy and hyperplasia with an increase in adipose tissue and free fatty acids resulting in endocrine and mechanical alteration. The latter is a factor causing dyspnea due to decreased lung volume and ventilator capacity associated with the increased abdominal pressure in the diaphragm [15]. High blood pressure (HBP) and insulin resistance or diabetes often accompanies obesity; therefore, obesity and its associated chronic conditions have more than one pathophysiological pathway of enabling severe manifestations in COVID-19 patients.

Even when asthma has not been identified as a strong risk factor for acquiring coronavirus disease, asthma patients, mainly those poorly controlled, could be at the highest risk of developing severe COVID-19 due to yet unknown immune networks that involve inflammatory cytokines [16]. Acute exacerbations of asthma require early recognition and intervention before a severe and life-threatening course of the condition [17]. Therefore, in the presence of COVID-19, people with asthma may require in-hospital assistance to the possible development of severe outcomes in the acute asthma exacerbations [18]. On the other hand, Chronic Obstructive Pulmonary Disease (COPD) may involve a combination of small airway disease and parenchymal destruction [19]. Therefore, acute exacerbations of COPD may as well derive to respiratory failure that may require treatment with mechanic ventilation in patients in the intensive care unit (ICU) [20]. The triage strategy for COVID-19 involves and defines the allocation of in-hospital services, intensive therapy management, and even mechanical ventilation with respirators. The triage and risk assessment tools have incorporated questionnaires that estimate the complication’s risks, age, gender, gestational status, lifestyle, and chronic comorbidities. Those include obesity, diabetes, hypertension, chronic kidney disease (CKD), cardiopathies, immune impairment, COPD, and asthma. Even when such indicators of complications are based on existing worldwide early evidence, assessing them is vital in clinical practice when deliberating to which patient should need the resource based on the odds of complications or even death.

In Mexico, the health system delivered self-applied risk assessment estimation tools and comprehensive triage guides with clinical practice algorithms. Both kinds of instruments are based on age and comorbidities, which are grouped by the affected system or organ (such as COPD and asthma, grouped as respiratory diseases). That practice may or may not be of independent prognostic importance for hospitalized patients with COVID-19 severe cases [21]. Therefore, analyzing the severity of the events that occurred in hospitalized patients with severe COVID-19 can help to identify the characteristics and comorbidities that represent increased odds for a poor outcome. The objective of the study is to analyze the underlying conditions of patients hospitalized for COVID-19 in Mexico concerning four severity outcomes (a) pneumonia, (b) mechanical ventilation (c) intensive care therapy, and (d) death.

## Materials and methods

This study is a retrospective cohort based on the epidemiologic case of all laboratory-confirmed patients with the COVID-19 infection that required hospitalization and complemented with the medical records of in-hospital assistance until discharge. In Mexico during the contingency, public hospitals in all 32 states were reconverted to receive and provide attention to COVID-19 patients specifically, and all hospitalizations derived from COVID-19 were notified mandatorily thru real-time epidemiologic surveillance system with the international classification of diseases under code U07.1. [22]. Ethical approval and Institutional Review Board for this study were exempted by Universidad Marista de Merida Board of Ethics because this study derives from Open Access anonymized dataset provided under the Mexican board of health through its General Directorate of Health Information webpage.

A standardized epidemiological case study form for each patient, including comorbid conditions, is generated, including clinical manifestations for triage. The in-hospital clinical course section is added if patients are hospitalized. Once updated, verified, and ratified, case studies are uploaded to the website of the department of health information as open access data sets. For the present study, we used the anonymized, open-access datasets. Data sets were coded, transformed into dependent and independent variables.

Independent variables were grouped as sociodemographic, comorbidities, and clinical. The dependent variables represented four possible severity outcomes that occurred during hospitalization including (a) pneumonia, (b) mechanical ventilation (c) intensive care unit, and (d) death; all of them were coded as binary. The frequencies, means, and percentages are presented and compared between groups for severity outcome distribution. The hospitalization rates are presented by 100,000 inhabitants, and for each severity outcome, percentages were calculated, while interstate variability was also assessed for each variable.

Severity outcomes were analyzed in separate binary logistic regression models adjusted by age and gender. For statistical significance, a p-value <0.05 was considered significant, while odds ratios were compared to 1.00 as a reference value, where significant values <1.00 were indicators as protective and >1.00 as of increased risk; finally, post hoc tests were performed to assess goodness of fit. All statistical analysis was performed using Stata 15 software.

## Results

To July 30, 2020, 69,334 patients with laboratory-confirmed SARS-CoV-2 infection were hospitalized in Mexican reconverted hospitals; male patients represented 62.61%, and the mean age was 55.29 years. Adjusted for age groups, Pediatric patients (age <15) represented 0.11% (N=804) and older adults (age >65) 27% (N=18,780) of hospitalized patients; median hospital stay was of five days. One-fourth of hospitalizations related to patients with obesity (23.56%), more than half of the patients had at least one previously diagnosed comorbidity. Diabetes was present among 30% of all hospitalized patients, hypertension was the most common comorbidity in 34% cases, cardiovascular diseases were present in 0.42%, CPK 0.5%, immune impairment 0.25%, and respiratory diseases 0.56%. Additionally, 0.82% were active smokers, and 1.7% (N=442) of hospitalized women were pregnant.

Around two thirds of patients developed pneumonia 66.5% (N=46,107); 9.6% (6,706) received mechanic ventilation; 8.56% (5,937) were located at the ICU and 35.72% (N=24,770) died. Regarding the interstate variability, the median rate of hospitalizations per 100,000 inhabitants was 39.7 (Range = 16.4-137.6); the median of pneumonia percentage among hospitalized patients was 63.6 (Range=42.1-79.6), the median of mechanical ventilation percentage was 8.8 (Range=3.3-14.9), the interstate median percentage for intensive care was 9.3 (Range=2.1-28.5), and the median percentage of in-hospital deaths was 33.5 (Range=18.6-47.7). (See supplemental material for maps describing severity outcome distribution in Mexican states).

Table 1 shows the distribution of comorbidities and severity outcomes by age groups of pediatric, young/adults, and the geriatric population. The differences showed how pneumonia affected more than 70% of geriatric patients, young and adult patients had a higher prevalence of obesity; pediatric patients were more commonly assisted at ICU. Death was prevalent in more than half of geriatric patients.

**Table 1.**
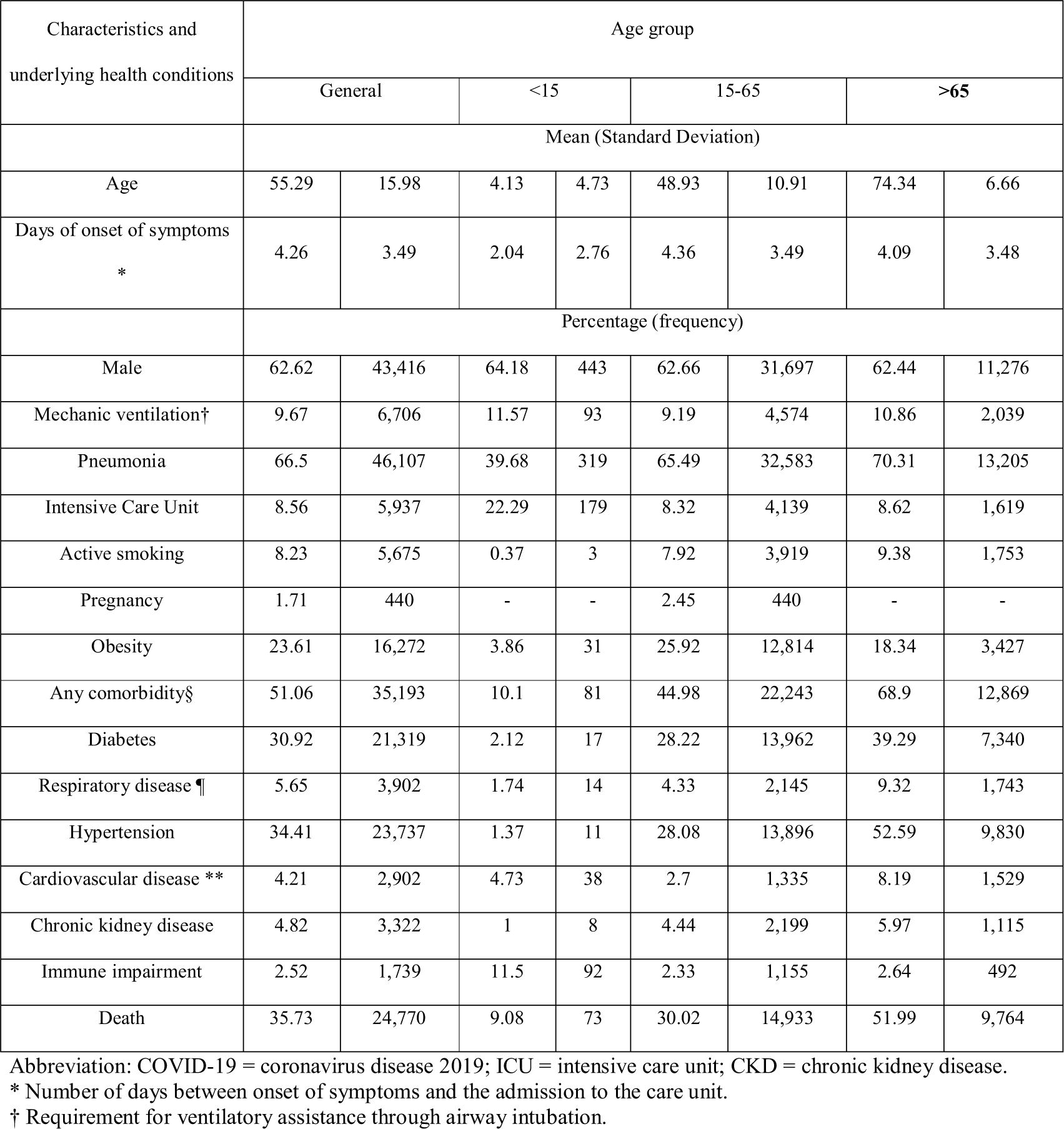
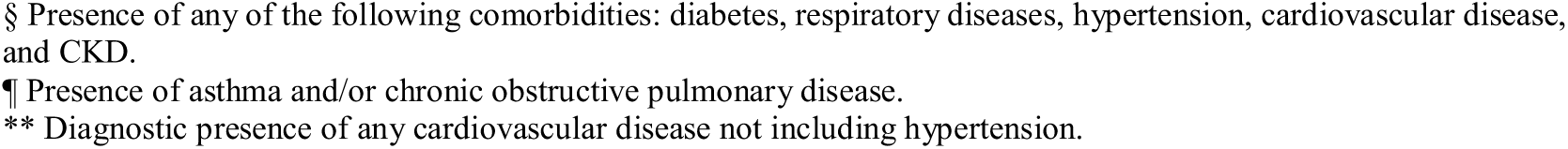
Characteristics underlying health conditions among hospitalized with COVID-19 by age group (N=69,334).

| Characteristics and<br>underlying health conditions | Age group |  |  |  |  |  |  |  |
| --- | --- | --- | --- | --- | --- | --- | --- | --- |
|  | General |  | <15 |  | 15-65 |  | >65 |  |
|  | Mean (Standard Deviation) |  |  |  |  |  |  |  |
| Age | 55.29 | 15.98 | 4.13 | 4.73 | 48.93 | 10.91 | 74.34 | 6.66 |
| Days of onset of symptoms<br>* | 4.26 | 3.49 | 2.04 | 2.76 | 4.36 | 3.49 | 4.09 | 3.48 |
|  | Percentage (frequency) |  |  |  |  |  |  |  |
| Male | 62.62 | 43,416 | 64.18 | 443 | 62.66 | 31,697 | 62.44 | 11,276 |
| Mechanic ventilation† | 9.67 | 6,706 | 11.57 | 93 | 9.19 | 4,574 | 10.86 | 2,039 |
| Pneumonia | 66.5 | 46,107 | 39.68 | 319 | 65.49 | 32,583 | 70.31 | 13,205 |
| Intensive Care Unit | 8.56 | 5,937 | 22.29 | 179 | 8.32 | 4,139 | 8.62 | 1,619 |
| Active smoking | 8.23 | 5,675 | 0.37 | 3 | 7.92 | 3,919 | 9.38 | 1,753 |
| Pregnancy | 1.71 | 440 | - | - | 2.45 | 440 | - | - |
| Obesity | 23.61 | 16,272 | 3.86 | 31 | 25.92 | 12,814 | 18.34 | 3,427 |
| Any comorbidity§ | 51.06 | 35,193 | 10.1 | 81 | 44.98 | 22,243 | 68.9 | 12,869 |
| Diabetes | 30.92 | 21,319 | 2.12 | 17 | 28.22 | 13,962 | 39.29 | 7,340 |
| Respiratory disease ¶ | 5.65 | 3,902 | 1.74 | 14 | 4.33 | 2,145 | 9.32 | 1,743 |
| Hypertension | 34.41 | 23,737 | 1.37 | 11 | 28.08 | 13,896 | 52.59 | 9,830 |
| Cardiovascular disease ** | 4.21 | 2,902 | 4.73 | 38 | 2.7 | 1,335 | 8.19 | 1,529 |
| Chronic kidney disease | 4.82 | 3,322 | 1 | 8 | 4.44 | 2,199 | 5.97 | 1,115 |
| Immune impairment | 2.52 | 1,739 | 11.5 | 92 | 2.33 | 1,155 | 2.64 | 492 |
| Death | 35.73 | 24,770 | 9.08 | 73 | 30.02 | 14,933 | 51.99 | 9,764 |

Abbreviation: COVID-19 = coronavirus disease 2019; ICU = intensive care unit; CKD = chronic kidney disease.
\* Number of days between onset of symptoms and the admission to the care unit.
† Requirement for ventilatory assistance through airway intubation.
§ Presence of any of the following comorbidities: diabetes, respiratory diseases, hypertension, cardiovascular disease, and CKD. ¶ Presence of asthma and/or chronic obstructive pulmonary disease. \*\* Diagnostic presence of any cardiovascular disease not including hypertension.

When comparing the proportional distribution of comorbidities among patients with or without severity outcomes (Table 2), it is noted that among patients who developed pneumonia, the percentage of active smokers, obesity prevalence, diabetes, hypertension, and cardiovascular diseases were higher than among those who did not develop pneumonia. Proportionally, among patients who needed mechanical ventilation during their hospitalization, pregnancy was less frequent, while obesity, diabetes, hypertension, and cardiovascular diseases were more frequent.

**Table 2.** Characteristics of comorbidities among COVID-19 hospitalized patients by severity outcomes (N=69,334).

| Variables | Pneumonia |  |  | Mechanical ventilation |  |  | Intensive Care |  |  | In-Hospital Death |  |  |
| --- | --- | --- | --- | --- | --- | --- | --- | --- | --- | --- | --- | --- |
|  | Yes | No | *p* | Yes | No | *P* | Yes | No | *p* | Yes | No | *p* |
| Active Smoking | **8.68** | 7.34 | **<0.001** | 8.7 | 8.18 | 0.072 | 8.17 | 8.24 | 0.43 | **8.87** | 7.87 | **<0.001** |
| Pregnancy | 1.05 | 2.89 | **<0.001** | 1.02 | **1.76** | **0.005** | 2.05 | 1.67 | 0.111 | 0.35 | **2.36** | **<0.001** |
| Obesity | **25.13** | 20.59 | **<0.001** | **28.69** | 23.07 | **<0.001** | **28.73** | 23.13 | **<0.001** | **24.74** | 22.98 | **<0.001** |
| Diabetes | **32.72** | 27.34 | **<0.001** | **35.72** | 30.41 | **<0.001** | **33.27** | 30.7 | **<0.001** | **37.33** | 27.36 | **<0.001** |
| Respiratory disease | 5.64 | 5.67 | 0.444 | 5.56 | 5.66 | 0.367 | 5.8 | 5.64 | 0.304 | **6.6** | 5.13 | **<0.001** |
| Hypertension | **35.72** | 31.82 | **<0.001** | **37.48** | 34.09 | **<0.001** | 34.32 | 34.42 | 0.437 | **42.5** | 29.93 | **<0.001** |
| Cardiovascular disease | **4.35** | 3.93 | **0.005** | **5.05** | 4.12 | **<0.001** | **4.8** | 4.15 | **0.009** | **5.4** | 3.55 | **<0.001** |
| Chronic kidney disease | 4.81 | 4.83 | 0.46 | 5.12 | 4.78 | 0.115 | **4.26** | 4.87 | **0.019** | **6.84** | 3.69 | **<0.001** |
| Immune impairment | 2.51 | 2.55 | 0.378 | 2.63 | 2.51 | 0.283 | **2.91** | 2.49 | **0.023** | **2.89** | 2.32 | **<0.001** |

\*Higher proportions of significant difference are presented in bold font.

In the group of patients who received treatment at the intensive care unit, the prevalence of smoking, obesity, cardiovascular diseases, CPK, and immune impairment were significantly higher than those who did not receive treatment at the ICU. Finally, the prevalence of pregnancy was lower in the group of patients who died. Still, the prevalence of all other comorbidities, including CPK and respiratory diseases (COPD and Asthma) were significantly higher.

After analyzing the association between the comorbidities and the odds for severity outcome occurrence (Table 3), it was found that the male gender and increasing age predicted every severity outcome. Pregnant women were significantly less prone to develop pneumonia and lower risk for in-hospital death. Diabetes and hypertension predicted every severity outcome significantly. Even when obesity increased the risk for pneumonia, more susceptible to receiving mechanical ventilation, and admitted at the ICU, obesity was not a predictor of in-hospital death.

**Table 3.**
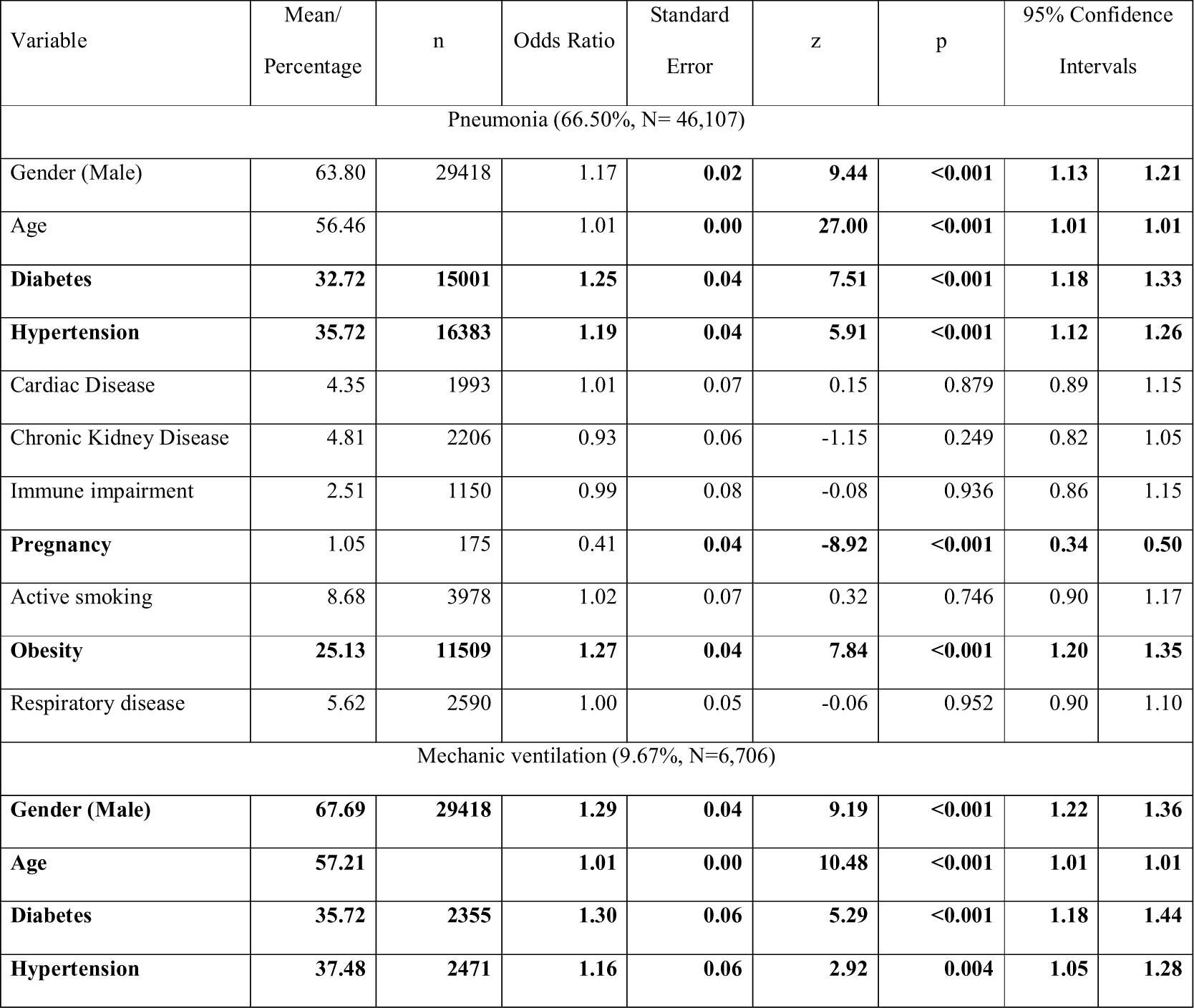
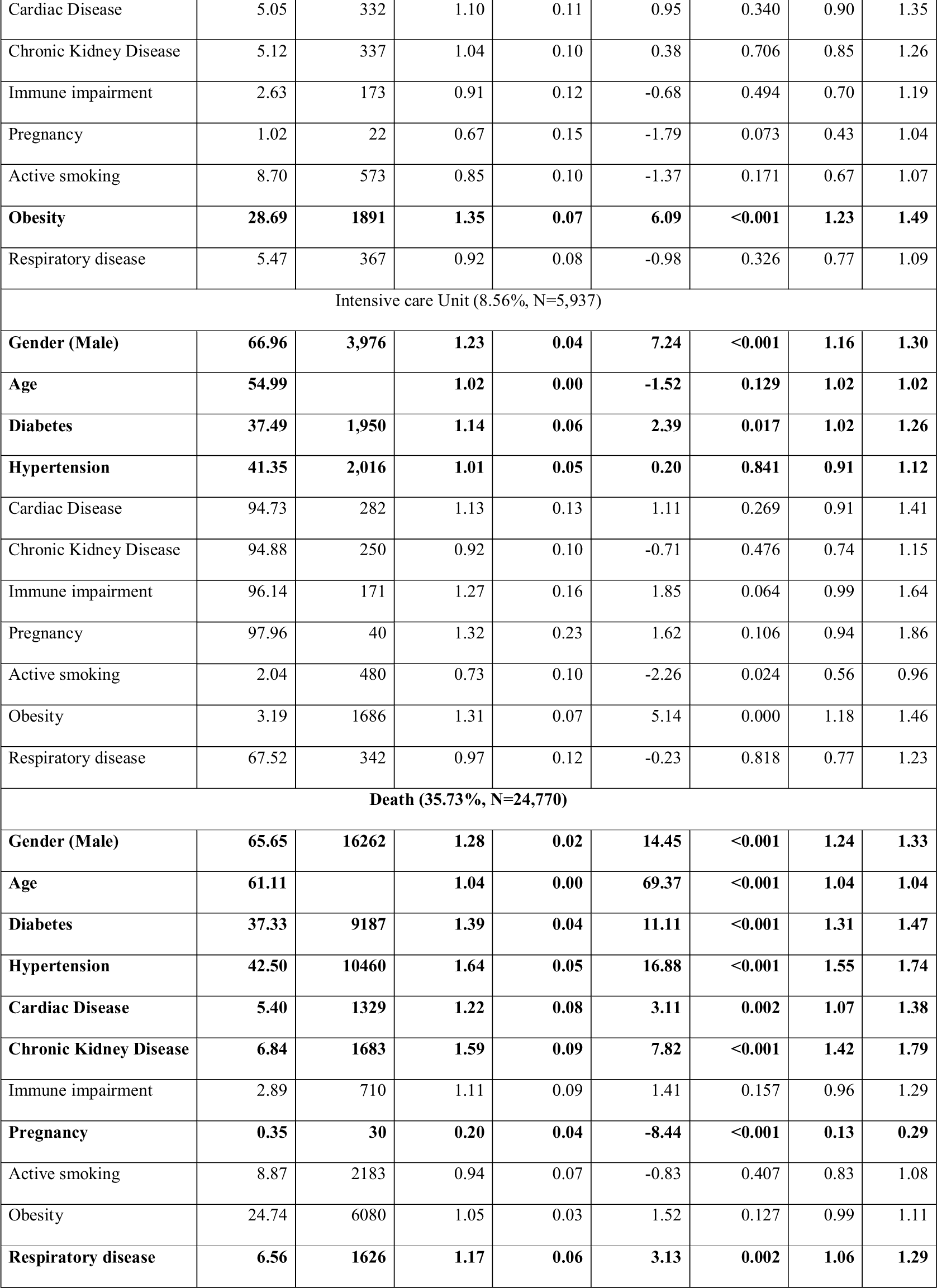
Association between comorbidities among hospitalized patients (N=69,334)

On the contrary, concerning respiratory diseases (asthma and COPD), patients were not more vulnerable to develop pneumonia. They were not prone to receive mechanical ventilation or assistance at the intensive care unit, but they were at higher risk of hospital death. Finally, *post hoc* analysis and likelihood in the regression model ensured goodness of fit with p=0.19.

## Discussion

The present study has provided an overview of the severity outcomes among hospitalized patients due to laboratory-confirmed COVID-19 infection in Mexico between February 28 and June 30, 2020. Nationwide, to the studied date, a hospitalization rate of 58.3 per 100,000 inhabitants was observed. By the end of April 2020, the in-hospital mortality rate was 27.85%. By July 30, we observed that 33.5% died while assisted in hospitals. The national in-hospital mortality rate increased during May and June.

The average age of the population of hospitalized patients in Mexico is between 55 years, with males’ predominance (62.62%). However, it changes concerning the population. In different studies, the median is at 41 years with a higher proportion in men (56.5%) and a wide range of patients who coexist with some medical comorbidity (12-67%), adding that the prevalence of hypertension (16%), diabetes (10.1%) and respiratory diseases (2%) in other populations was lower in comparison to this study [23]. In a follow-up and surveillance of cases study in the United States, the median was 48 years with similar proportions between the genders, thus indicating that the frequencies of comorbidities were different in contrast to this study, since there is a lower rate of patients with diabetes (30.2%) and cardiovascular diseases including hypertension (32.2%) in this population. However, the percentages of patients with respiratory diseases (17.5%), CPK (7.6%), and immunocompromised (5.3%) were higher in comparison to the hospitalized patients in the present study [5].

Concerning the pediatric patients, the average age in our population varies in comparison to other studies that had reported six years as the median age of pediatric patients. However, this differs according to the age range, determined to consider a pediatric patient in systematic reviews [24, 25]. Patel NA reported in a systematic review that the most common underlying clinical conditions in pediatric patients with COVID-19 disease were asthma, immunosuppression, and cardiovascular diseases, this conditions being prevalent in 21% of the patients, however, in our study, only 10% of patients had any comorbidity, adding that there were also cases of patients with diabetes, hypertension and CKD [26]. Concerning mechanical ventilation and ICU admission, the evidence reported that 0.7% of patients avoided both, which is lower than what we found in this study, with 11.57% of patients requiring mechanical ventilator assistance and 22.29% of patients admitted to the ICU. Even when we observed lower mortality (9.08%) among pediatric patients, compared to other groups of patients distributed by age, the pediatric mortality is almost ten times greater than the one reported by Meena J in 4,476 patients, of which five individuals died [27].

In the present study, patients aged >65, had a proportion of death (51.99%) that surpassed the frequency of survival when hospitalized. It is not surprising that all comorbidities were more prevalent among male elderly patients and that every severity outcome was more frequent. Odds of developing pneumonia, receiving mechanical ventilation, being assisted at the ICU, and having a fatal outcome during hospital stay were higher among men and tended to increase with age. These findings are consistent with similar clinical series from China [23, 28]. Still, they are discordant with the conclusions of the Martín-Sanchez, et al. study [29], where advanced age predicted not being admitted at the ICU. These findings from the Spain series may also have underlying explanations from non-clinical nature, partially derived from resource administration and hospital decision-making.

Comorbidities distribution among patients in this study is congruent with the high prevalence of chronic diseases among adult Mexican men, which reached 75.2% of the adult population according to the latest national health survey; additionally, 10.3% of the population has diabetes [30]. Mexico has been identified as the second country with the largest population affected by obesity [31].

Yang J et al., in their systematic review noted that hypertension and diabetes were the most common comorbidities followed by cardiovascular and respiratory diseases; they also mention that the severity and clinical course of COVID-19 disease were related to these health conditions. Furthermore, Du RH et al. indicate that patients over 65 have a higher risk of dying from the disease [7,32]. In the present study, hypertension and diabetes were also the most common comorbidities among hospitalized patients, and consistent with our results, the advanced age and those with pre-existing cardiovascular disease, were significant predictions of hospital mortality.

A study by Vardabas et al. reports that smoking is also associated with a negative progression of the disease since it presents a more significant probability (RR 1.4) of showing worsening of the symptoms to 2.4 times the likelihood of requiring mechanical ventilation, admission to the ICU, and risk of death [6]. However, in this study, that was not the case since smoking was not found to be a risk factor for severity outcomes.

We have presented how obesity was related to higher odds of developing pneumonia, mechanical ventilation, and assisted in the ICU. Still, we found that obesity was not a predictor of death. To some degree, our findings are similar to those reported by Tamara A, et al. where it was found that obesity predicted mechanical ventilation and that a higher degree of obesity was a more reliable predictor [33]. Mexican triage guidelines and official national algorithms for clinical practice describe the route of action and clinical care peculiarities of patients with pneumonia, diabetes, immune impairment, and CPK, consisting of individualized, isolated, or ICU management [21].

Based on our findings, we believe that these algorithms may have had a positive impact on patients’ decision-making with the mentioned comorbidities. Still, we consider that updates to such guidelines, including specific evaluation for asthma and COPD ungrouped, could benefit Mexican patients. Our findings showed that patients with respiratory conditions did not proportionally receive more frequently mechanical ventilation or admittance to intensive care unit even when they were significantly more susceptible to die, even after adjusting for age and gender.

### Limitations

Limitations of the present study derive from its retrospective design: first are those resulting from data obtained from the epidemiologic surveillance system Open Access datasets that may or may not include reporting or coding defects. Nevertheless, all information is validated and ratified at different administrative levels. The study does not allow us to analyze any acute comorbidities that could have happened simultaneously in hospitalized patients. Lastly, as in any other health system with COVID-19 sentinel surveillance, we may have unintentionally excluded patients hospitalized but were not tested for COVID-19.

### Implications and recommendations

Clinical practice guidelines, triage instruments, and algorithms have been precise in guiding hospital care for patients with pneumonia, diabetes, immune impairment, and CPK. However, they could still improve hospital assistance and define resource allocation for patients with algorithms for respiratory conditions, including asthma and COPD.

## Conclusions

Overall, in-hospital mortality in Mexico due to COVID-19 up to June 30 was 35.72%; among pediatric patients, it was 9.11%, and more than half of patients aged >65 died. The most common comorbidities were hypertension and diabetes. Male gender, age, hypertension, and diabetes predicted pneumonia, mechanical ventilation, attention in ICU, and death. Obesity increased the odds of pneumonia, of receiving mechanical ventilation and admittance at the ICU, but did not increase death risk.

Contrastingly, patients with CPOD and/or asthma had increased risk for death but did not receive mechanical ventilation nor were assisted at the intensive care unit more frequency.

## Data Availability

The dataset corresponds to open access, anonymized and under Creative Commons license Available at https://datos.gob.mx/
For our study's purpose, we generated variables and transformed information into epidemiologic measures.
Spreadsheets are downloaded initially with the CSV extension and were converted to Excel and Stata formats by authors.

## Author Contributions

G.C., N.M.-D., A.A.-B. Contributed to the conception and design of the manuscript; K.D.S.-Z., A.R.-P., M.A.-M., and O.C.-K. contributed to collection and management of the hospitalizations and the mortality datasets. N.M.-D. and A.A.-B. performed data analysis and interpretation. All authors drafted and revised the manuscript. All authors read and approved the final manuscript and take full responsibility for the manuscript.

